# A Machine Learning Approach to Predicting Heterogeneous Lipid Responses to Statins in Type 2 Diabetes

**DOI:** 10.64898/2026.01.01.26343317

**Authors:** Carl Harris, Daniel Olshvang, Rama Chellappa, Prasanna Santhanam

## Abstract

**Background:** Inter-individual variability in lipid response to statin therapy poses a major challenge in cardiovascular risk reduction, particularly among patients with type 2 diabetes mellitus (T2DM), who exhibit complex dyslipidemia and elevated cardiovascular risk. While randomized trials provide population-level estimates of treatment efficacy, individualized prediction of lipid changes, and corresponding uncertainty, remains limited. Using rich clinical data from the Action to Control Cardiovascular Risk in Diabetes (ACCORD) study, this work sought to quantify and predict person-level changes in multiple lipid fractions following statin initiation.

**Methods:** Among 3,509 ACCORD participants with T2DM who were newly initiated on simvastatin and had 12-month lipid follow-up, we developed machine learning models to predict changes in LDL-C, HDL-C, triglycerides, VLDL, and total cholesterol using only baseline clinical features. Five model classes were evaluated using nested cross-validation; Lasso regression emerged as the best-performing model for LDL-C. To provide clinically interpretable and statistically valid uncertainty estimates, we applied distribution-free conformal prediction to generate individual-level prediction intervals at multiple confidence levels. Feature importance was quantified using SHAP values.

**Results:** Longitudinal lipid responses showed substantial heterogeneity across individuals. Predictability varied markedly by lipid fraction: LDL-C (𝑅^2^ = 0.52; 𝑀𝐴𝐸 = 18.3 𝑚𝑔/𝑑𝐿) and total cholesterol (𝑅^2^ = 0.41) were moderately estimable from baseline data, whereas HDL-C (𝑅^2^≈ 0.04) exhibited minimal predictability, and triglycerides and VLDL demonstrated intermediate but volatile behavior. Baseline lipid values were the strongest predictors across all outcomes, followed by markers of statin exposure and metabolic status. Conformal prediction achieved accurate empirical coverage across all lipid fractions (≈ 90% at nominal 90% confidence), with interval widths scaling appropriately to biological variability. Undercoverage occurred primarily among individuals predicted to have minimal lipid changes.

**Conclusions:** Machine learning models can meaningfully predict LDL-C and total cholesterol responses following statin initiation in patients with T2DM, while HDL-C and triglyceride responses remain dominated by unmeasured or stochastic factors. Conformal prediction adds a crucial layer of reliability by providing calibrated, patient-specific uncertainty intervals, supporting safer and more transparent clinical decision-making. These findings highlight both the promise and the current limitations of EHR-based precision lipid management and underscore the need to incorporate genetic, metabolic, and behavioral data to capture the full spectrum of lipid response variability.

## Introduction

Lowering atherogenic lipoproteins with statins is a cornerstone of cardiovascular risk reduction, yet the lipid response to treatment varies markedly between individuals^1^. In routine practice, some patients experience large low-density lipoprotein cholesterol (LDL-C) and total cholesterol (TC) reductions while others show modest changes, and high-density lipoprotein cholesterol (HDL-C) and triglyceride (TG) responses are similarly heterogeneous^2^. In routine clinical practice, this variability means that some patients experience large LDL-C reductions while others show only modest changes, even when standardized for dose and potency^3^. This variability is multifactorial, reflecting a complex interplay of baseline lipid levels, adherence to medication, concomitant drug therapies, pre-existing metabolic comorbidities, and underlying biological dynamics, including factors like genetics (e.g., polymorphisms in lipid metabolism genes such as *SLCO1B1*^4^ and *HMGCR*^5^) and gut microbiome composition^6^. Recent multi-omic studies and reviews underscore the substantial contribution of these factors to the on-target statin effects on LDL-C^7^.

The complexity of heterogeneous lipid response is magnified significantly in patients with Type 2 Diabetes Mellitus (T2DM). This population is characterized by significantly elevated cardiovascular risk and challenging dyslipidemic phenotype, often defined by elevated TG, reduced HDL-C, and a predominance of small, dense atherogenic LDL-C particles^8^. The Action to Control Cardiovascular Risk in Diabetes (ACCORD) trial^9^ provides a unique platform to examine individual-level lipid responses to statins in adults with T2DM^10^. ACCORD enrolled over 10,000 participants (ages 40 to 79) with T2DM and a history of cardiovascular disease (CVD) or multiple CVD risk factors across 77 clinical centers in North America and randomized them to intensive versus standard strategies for glycemic, blood pressure, and lipid control. The lipid trial component specifically compared a combination therapy (fenofibrate plus simvastatin) versus simvastatin alone, while all participants in the main trial had standardized access to statins as part of cardiovascular risk management. The combination was selected because fenofibrate and simvastatin target different aspects of dyslipidemia, with fenofibrate primarily lowers triglycerides and raises HDL, whereas simvastatin reduces LDL cholesterol^11^, leading to the hypothesis that dual therapy might improve cardiovascular outcomes in high-risk patients. Detailed longitudinal data, including medication use, laboratory measurements, and clinical outcomes, enable precise modeling of changes in LDL-C, HDL-C, and TG in the context of statin therapy^12,13^.

The inability to accurately predict an individual patient’s magnitude of response to statin initiation or modification translates directly into residual cardiovascular risk, necessitates a move beyond population-averaged efficacy measures derived from randomized controlled trials (RCTs) toward personalized, precision prediction. While RCTs confirm the average benefit of statin therapy^14,15^, the crucial clinical requirement is to identify the *individual* patient who will experience an inadequate therapeutic reduction (a “non-responder”) or an exaggerated response, thereby facilitating individualized therapeutic targeting and the assessment of residual CV risk in patients.

Modern machine learning (ML) methods offer a powerful opportunity to leverage rich clinical data to estimate patient-specific changes in LDL-C, HDL-C, and TG following statin initiation or modification^16,17^. Unlike conventional linear regression approaches, ML models can automatically capture high-order nonlinearities and interactions among key covariates such as glycemic control, renal function, body weight, and concomitant therapies, enabling a more faithful representation of the complex biological pathways influencing lipid metabolism in diabetes^18^.

In safety-critical domains such as personalized drug dosing and CV medicine, a prediction is not fully actionable unless accompanied by a measure of confidence. ML models, which primarily exploit correlations in data rather than guaranteed causal links, require a rigorous quantification of their predictive error to prevent dangerous over-reliance in complex cases^19^. Relying solely on a point estimate, even one with high average accuracy, can be misleading^20^, particularly when the underlying biological process is inherently noisy or volatile, such as in triglyceride metabolism. To meet the high clinical standard for reliability, this study employed Conformal Prediction (CP), a powerful, distribution-free, and model-agnostic framework for uncertainty quantification^21^. CP provides statistically valid prediction intervals that contain the true outcome value with a specified probability (e.g., 90% or 95%), irrespective of the underlying data distribution or modeling assumptions^22^. The application of CP generates calibration-guaranteed prediction intervals. This framework provides clinicians with directly interpretable confidence ranges (e.g., “The patient’s LDL-C change will fall between 𝑋 and 𝑌 mg/dL with 𝑍% confidence”).

Although recent ML-based efforts have explored the prediction of LDL-C goal attainment and statin response in diverse population cohorts^23–26^, few have focused specifically on individuals with diabetes or attempted to model multiple lipid fractions simultaneously, which is an essential step toward understanding the multidimensional dyslipidemia characteristic of this high-risk group. Further, none have considered uncertainty quantification in their methodologies. In this study, we sought to quantify and predict the heterogeneous longitudinal lipid responses to statin therapy in patients with T2DM using clinical data from the ACCORD trial. We begin with a binary classification model trained to differential LDL levels at a threshold of 70 mg/dL and 100 mg/dL. Then, by applying a regression model and conformal prediction, we aimed to generate individualized estimates of changes in LDL-C, HDL-C, TG, VLDL, and TC between baseline and a one-year follow-up, along with rigorous, calibration-guaranteed uncertainty intervals for each.

## Methods

### Study population

The study population consisted of 10,251 participants with T2DM enrolled in the ACCORD trial. Of these, 5,514 individuals participated in the ACCORD lipid trial arm, which compared combination therapy with fenofibrate plus simvastatin versus simvastatin alone. Within the lipid arm, 3,723 participants had no recorded simvastatin dose at baseline but were subsequently prescribed simvastatin as part of trial-directed lipid management. Among this subgroup, 3,509 participants had complete 12-month lipid panel follow-up data and were included in the primary longitudinal lipid response analyses. A description of patient characteristics is shown in **Table 1**.

**Table 1.** Description of sample population. Shown are demographics, clinical variables, and common medication use for the overall ACCORD cohort and for the study cohort used in this analysis following sample selection. Abbreviations: UACR (urine albumin-to-creatinine ratio); ACE (angiotensin-converting enzyme).

|  | ACCORD Cohort | Study Cohort |
| --- | --- | --- |
| Patients (*n*) | 10,251 | 3,509 |
| Age (mean ± std.) | 62.8 ± 6.6 | 62.5 ± 6.4 |
| Female gender (%) | 3,952 (38.6 %) | 1,187 (33.8%) |
| Race and ethnicity |  |  |
| White | 6,393 (62.4%) | 2,359 (67.2 %) |
| Black | 1,953 (19.1%) | 437 (12.5 %) |
| Hispanic/Latino | 737 (7.2%) | 229 (6.5 %) |
| Other | 1,168 (11.4%) | 484 (13.8 %) |
| Clinical history |  |  |
| History of cardiovascular disease | 3,609 (35.2%) | 1,130 (32.2 %) |
| Lipid profile |  |  |
| Total cholesterol, mg/dL | 183.3 ± 41.7 | 181.0 ± 38.5 |
| LDL cholesterol, mg/dL | 104.9 ± 33.8 | 105.1 ± 31.7 |
| HDL cholesterol, mg/dL | 41.9 ± 11.6 | 38.1 ± 7.8 |
| Triglycerides, mg/dL | 190.1 ± 148.0 | 194.2 ± 116.6 |
| VLDL cholesterol, mg/dL | 36.5 ± 24.3 | 37.7 ± 20.5 |
| Other clinical variables |  |  |
| Fasting plasma glucose, mg/dL | 175.2 ± 56.0 | 176.7 ± 54.4 |
| eGFR, ml/min/1.73 m<sup>2</sup> | 91.1 ± 27.1 | 90.3 ± 23.6 |
| UACR, mg/g (IQR) | 15.0 (7.0, 57.0) | 14.0 (7.0, 54.5) |
| Serum creatinine, mg/dL | 0.91 ± 0.23 | 0.91 ± 0.22 |
| Serum potassium, mEq/L | 4.47 ± 0.47 | 4.47 ± 0.42 |
| Medication use |  |  |
| ACE inhibitor | 5,580 (54.5%) | 1,789 (51.0 %) |
| Biguanide (metformin) | 6,554 (63.9%) | 2,295 (65.4 %) |
| Aspirin | 5,597 (54.6%) | 1,901 (54.2 %) |

### Preprocessing

#### Baseline data

The primary data sources for our analysis included: the ACCORD key file containing participant demographics and randomization information (9 variables; demographics, randomization information, history of CVD), concomitant medications data (54 categories), lipid laboratory values (5 variables; TC, TG, LDL-C, HDL-C, VLDL), other laboratory results (9 variables; e.g., fasting plasma glucose, serum creatinine, eGFR), blood pressure (SBP, DBP, HR), lipid medication management data (4 variables; simvastatin dose on baseline visit entry/exit, medication adherence, and other lipid-modulating medication), and numerical values from a baseline physical exam history (57 items).

The ACCORD key file was processed to extract demographic information. Race/ethnicity was converted from categorical format to binary indicator variables (White, Black, Hispanic, Other) using one-hot encoding. Concomitant medication data were extracted for the baseline visit only. This dataset contained binary indicators for numerous medication classes including antihypertensive medications, diabetes medications, lipid-lowering medications, and other medications (aspirin, anticoagulants, antidepressants, etc.). Laboratory data were extracted for the baseline visit. This included lipid panel measurements, metabolic parameters, and renal function markers. A subset of numerically-encoded variables from the baseline history and physical examination were selected, including social history, medical history (e.g., years with diabetes/dyslipidemia/hypertension, CVD history, neuropathy, depression, eye disease, etc.), physical examination findings, and insurance/medication coverage information. Lipid medication management data were processed to extract statin dosing information at the start and end of the baseline visit for the lipid intervention arm (*n = 5,514*). Only participants not taking simvastatin at the start of their baseline visit but prescribed it following the visit (*n = 3,723*) and with a lipid panel after 12 months (*n = 3,509*) were included in the study. All baseline features were merged using the participant identifier (*MaskID*).

Given the presence of missing values across multiple variables in the combined baseline dataset, an iterative imputation approach was employed. We selected multiple imputation by chained equations^27^, which models each feature with missing values as a function of other features in a round-robin fashion and preserves relationships between variables making it suitable for datasets with complex interdependencies, as in our case. The imputation process was run for 10 iterations, reaching convergence.

#### Longitudinal analyses

For our primary analyses, we were interested in the change in lipid levels over the 12-month period following the baseline visit. Specifically, given a lipid value 𝐿𝑉 (e.g., LDL-C or HDL-C), our outcome variable of interest is:

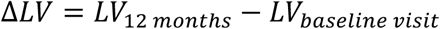

Then, we attempted to predict the change in each lipid fraction in the lipid panel using only the measurements and data available at baseline as features. Essentially, the goal was to identify whether it was possible to predict deviations from a patient’s “set point” lipid value (defined as the baseline value) following intervention and conditioned on observable covariates (e.g., demographics, baseline laboratory values, etc.). The primary outcome of interest was the change in LDL-C from baseline to 12-month follow-up. Secondary analyses examined changes in other lipid parameters including TC, HDL-C, TG, and VLDL using the same approach.

### Estimation of lipid values

#### Model selection and training

We evaluated five machine learning models to evaluate our approach. Ridge regression extends linear regression by incorporating L2 regularization to reduce overfitting and handle multicollinearity^28^. Lasso Regression employs L1 regularization, which prevents overfitting and performs automatic feature selection by driving coefficients of less important features to zero^29^. ElasticNet combines both L1 and L2 regularization penalties, offering a balance between Ridge and Lasso that can handle correlated features while still performing feature selection^30^. Random Forest (RF) is an ensemble method that aggregates predictions from multiple decision trees, generating final predictions via bootstrap aggregation and feature randomization^31^. Finally, XGBoost (Extreme Gradient Boosting) is a gradient boosting framework that sequentially builds an ensemble of weak learners, optimizing a differentiable loss function through gradient descent, and incorporates regularization techniques to prevent^32^. Each model was trained to minimize the mean-squared error between predicted and true values.

Hyperparameter optimization was performed to prevent overfitting, using nested cross-validation and ensure unbiased performance estimates. The outer loop employed 5-fold cross-validation, while the inner loop utilized 5-fold cross-validation. Within each outer fold, hyperparameters were optimized using randomized search with 100 iterations, systematically exploring the hyperparameter space defined for each model type (see **Table S2** for detailed hyperparameter ranges). The optimal hyperparameter configuration for each model was selected as the most frequently chosen combination across all outer folds. Model evaluation was subsequently conducted using 10-fold cross-validation with the selected hyperparameters. Performance was assessed using the coefficient of determination (𝑅^2^), which quantifies the proportion of variance in the outcome explained by the model (on the range from 0 to 1), and mean absolute error (MAE), which is the average prediction error magnitude:

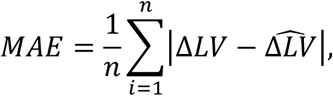

Where 𝑛 is the number of observations and Δ𝐿𝑉 and Δ𝐿̂𝑉 are the actual and predicted lipid values of interest.

#### Feature importance analysis

To interpret model behavior and identify the baseline clinical variables most strongly associated with lipid response trajectories, we conducted a post-hoc feature importance analysis using SHAP (SHapley Additive exPlanations)^33^. SHAP provides a unified, model-agnostic framework for quantifying each feature’s marginal contribution to an individual prediction by computing Shapley values, which is a game-theoretic quantification of credit among inputs in a competitive game.

This approach is particularly suited for complex, potentially nonlinear models, as it enables consistent comparison of feature effects even when predictors are correlated or have heterogeneous scales. We computed SHAP values on the final cross-validated model for each lipid outcome. For each lipid fraction, SHAP values were calculated for all participants in the held-out folds and then aggregated to derive global feature importance scores, defined as the mean absolute Shapley value across all observations. Features were subsequently ranked to identify the most influential predictors for each outcome.

### Uncertainty quantification

To provide clinically meaningful prediction intervals with guaranteed coverage properties, we employed conformal prediction, a distribution-free framework for uncertainty quantification. Conformal prediction provides statistically valid prediction intervals that are guaranteed to contain the true outcome value with a specified probability (e.g., 90% or 95%), regardless of the underlying data distribution or model assumptions.

The conformal prediction approach was implemented using the Model-Agnostic Prediction Intervals (MAPIE) library^34^ with the “plus” method. This method works by: (1) training the model on the proper training set; (2) computing nonconformity scores (prediction errors) on the calibration set; (3) determining the appropriate quantile of these scores to construct prediction intervals; and (4) applying these intervals to test set predictions. The prediction intervals have the property that, for a specified confidence level 𝛼 (e.g., 𝛼 = 0.1 for 90% confidence), at least (1 − 𝛼)% of true values will fall within the predicted intervals, providing a distribution-free guarantee of coverage.

We evaluated conformal prediction intervals at multiple confidence levels (80%, 90%, 95%, 99%) and assessed empirical coverage (the proportion of test samples where true values fell within predicted intervals) to verify that the method achieved the target coverage rates. Additionally, we examined interval widths to assess the precision of uncertainty estimates, with narrower intervals indicating more precise predictions.

## Results

### Baseline Characteristics of the Study Population

The final analytical cohort consisted of 3,509 participants with T2DM from the ACCORD study. The mean age was 62.5 ± 6.4 years, and 33.8% (*n = 1,187*) were female. The population was racially diverse, identifying predominantly as White (67.2%) or Black (12.5%). At baseline, participants exhibited a mean HbA1c (derived from fasting glucose surrogates) consistent with established diabetes. The mean baseline LDL cholesterol (LDL-C) was 105.1 ± 31.7 mg/dL, while triglycerides (TG) averaged 194.2 ± 116.6 mg/dL.

### Prediction of Categorical Lipid Changes

Prediction performance for categorical LDL-C target attainment of < 70 and < 100 mg/dL at 12 months is shown in **Figure 1**. The model demonstrated moderate discriminatory ability for both guideline thresholds, with slightly stronger performance for the more stringent target. For attainment of LDL-C < 70 mg/dL, the model achieved an AU-ROC of 0.745 and an AU-PRC of 0.486, substantially exceeding the baseline precision corresponding to outcome prevalence (AP = 0.238). For the LDL-C < 100 mg/dL threshold, discrimination was modestly lower by AU-ROC (0.712) but markedly higher in precision-recall space, with an AU-PRC of 0.838 compared with a baseline AP of 0.693. These results indicate that while both targets are predictably identifiable from baseline clinical data, prediction of the more prevalent < 100 mg/dL outcome yields higher precision across recall levels, whereas prediction of < 70 mg/dL reflects greater class imbalance and biological difficulty despite comparable overall discrimination.

**Figure 1.**
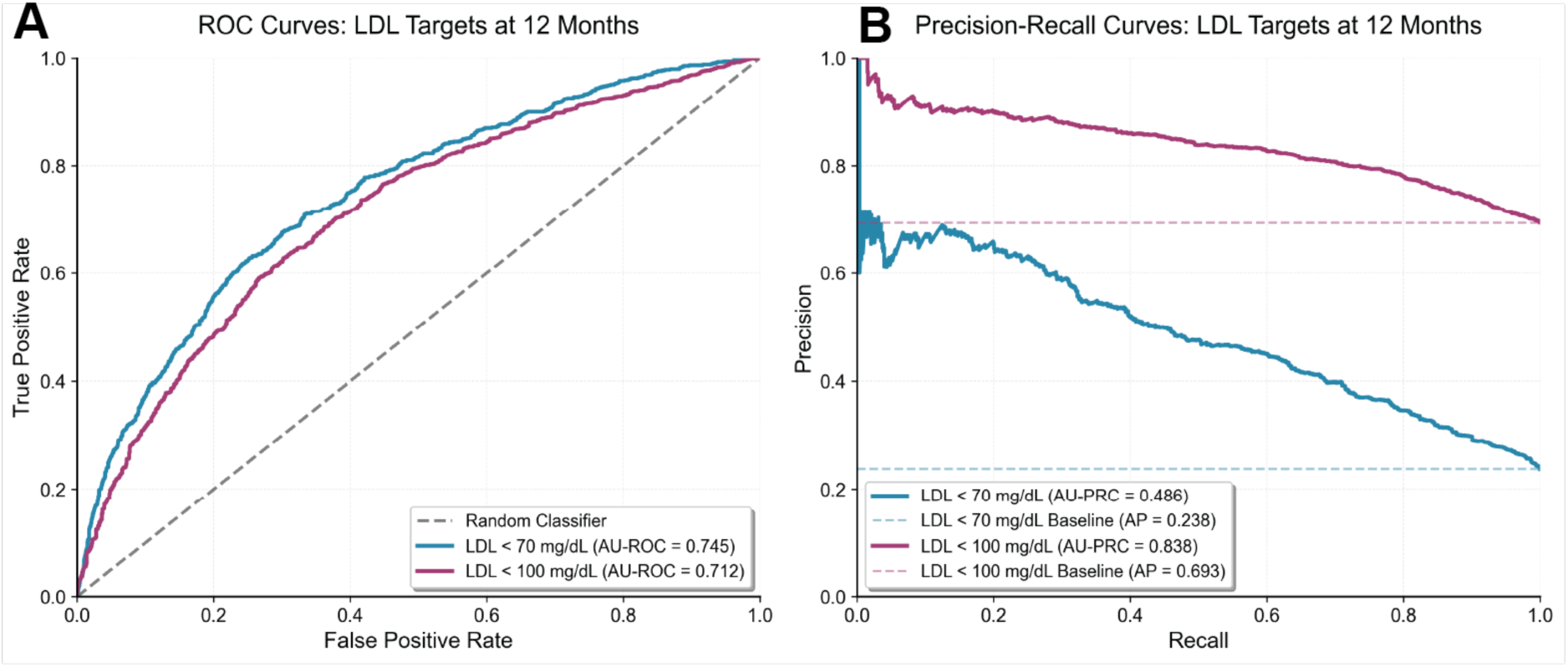
Discriminative performance in binary classification task. Plot shows performance of the baseline clinical model for predicting LDL-C target attainment at 12 months following statin initiation in patients with T2DM. **(A)** Receiver operating characteristic (ROC) and **(B)** precision-recall (PR) curves are shown for LDL-C thresholds of < 70 mg/dL and < 100 mg/dL.

### Prediction of Longitudinal Lipid Changes

We began by estimating changes in all lipid parameters from baseline after 12-month follow-up across our five models of consideration. We selected *Lasso Regression* as our base model because it obtained the highest performance on the LDL-C estimation task (see **Table S1**).

The model demonstrated the strongest predictive performance for LDL-C and Total Cholesterol. For the primary outcome of LDL-C change, the model achieved a coefficient of determination (𝑅^2^) of 0.520 across test folds, with a Mean Absolute Error (MAE) of 18.29 mg/dL. This indicates that approximately 52% of the variance in LDL response can be explained by baseline clinical features. Similarly, TC showed robust predictability (𝑅^2^= 0.411; MAE = 23.19 mg/dL; see **Figure S4**).

In contrast, the model was less effective at predicting changes in HDL-C (𝑅^2^ = 0.040; MAE = 5.14 mg/dL), suggesting that HDL response trajectories are dominated by stochastic factors or biological variables not captured in standard clinical data. Triglycerides (𝑅^2^ = 0.188; MAE = 63.99 mg/dL) and VLDL (𝑅^2^ = 0.171; MAE = 11.79 mg/dL) showed intermediate predictability, reflecting the high biological variability and wide dynamic range associated with these lipid fractions (see **Table 2**).

**Table 2.** Prediction performances using Lasso Regression.

| Lipid fraction | Test $R^2$ | MAE (mg/dL) | Empirical coverage (%) | Mean Interval width (mg/dL) |
| --- | --- | --- | --- | --- |
| LDL-C | 0.520 | 18.29 | 90.9 | 75.6 |
| HDL-C | 0.040 | 5.14 | 90.3 | 21.7 |
| Triglycerides | 0.188 | 63.99 | 90.8 | 257.5 |
| VLDL | 0.171 | 11.79 | 90.7 | 48.1 |
| Total Cholesterol | 0.411 | 23.19 | 90.5 | 92.3 |

As shown in **Figure 2**, the predicted versus actual plot shows that model estimates for LDL-C cluster closely around the identity line, indicating strong agreement between predicted and observed changes. In contrast, TG and VLDL display markedly wider scatter, reflecting their high biological volatility and the model’s limited ability to capture their large dynamic range. HDL-C exhibits similarly poor alignment, with points diffusely distributed around a predicted HDL change of 0, indicating poor discriminative capacity by the model.

**Figure 2.**
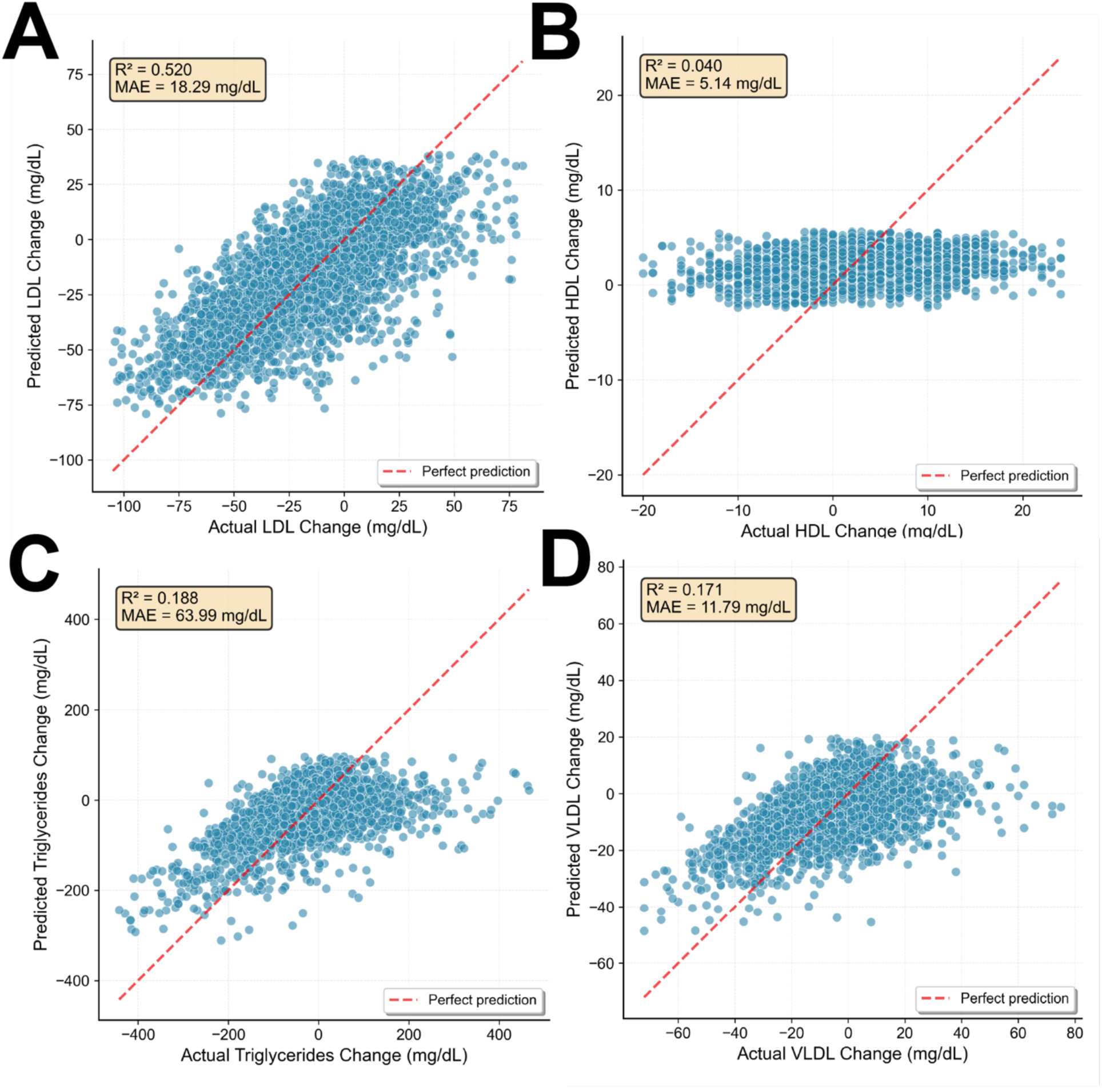
Comparison between actual and predicted lipid values. Plotted are correlation scatter plots for changes in **(A)** LDL-C, **(B)** HDL-C, **(C)** TG, and **(D)** VLDL. For visual simplicity, extreme values (those beyond 3 SD of the mean) are not plotted (but are included in the 𝑅^2^calculation).

### Key Determinants of Lipid Response

To interpret the drivers of the model’s predictions, we analyzed SHAP values (**Figure 3**). Across all lipid fractions, the baseline value of the specific lipid being predicted was consistently the strongest feature, indicating that the magnitude of change is heavily dependent on the starting point (likely capturing both regression to the mean and physiological limits of lipid modification). Beyond baseline lipid levels, several clinical and pharmacologic variables contributed meaningfully to predictions. Indicators of statin exposure ranked among the next most influential factors for LDL-C and TC. Collectively, these patterns suggest that while certain lipid responses (most notably LDL-C) are driven by well-captured baseline physiology and treatment intensity, others remain influenced by unmeasured behavioral, genetic, or metabolic factors not reflected in standard clinical data.

**Figure 3.**
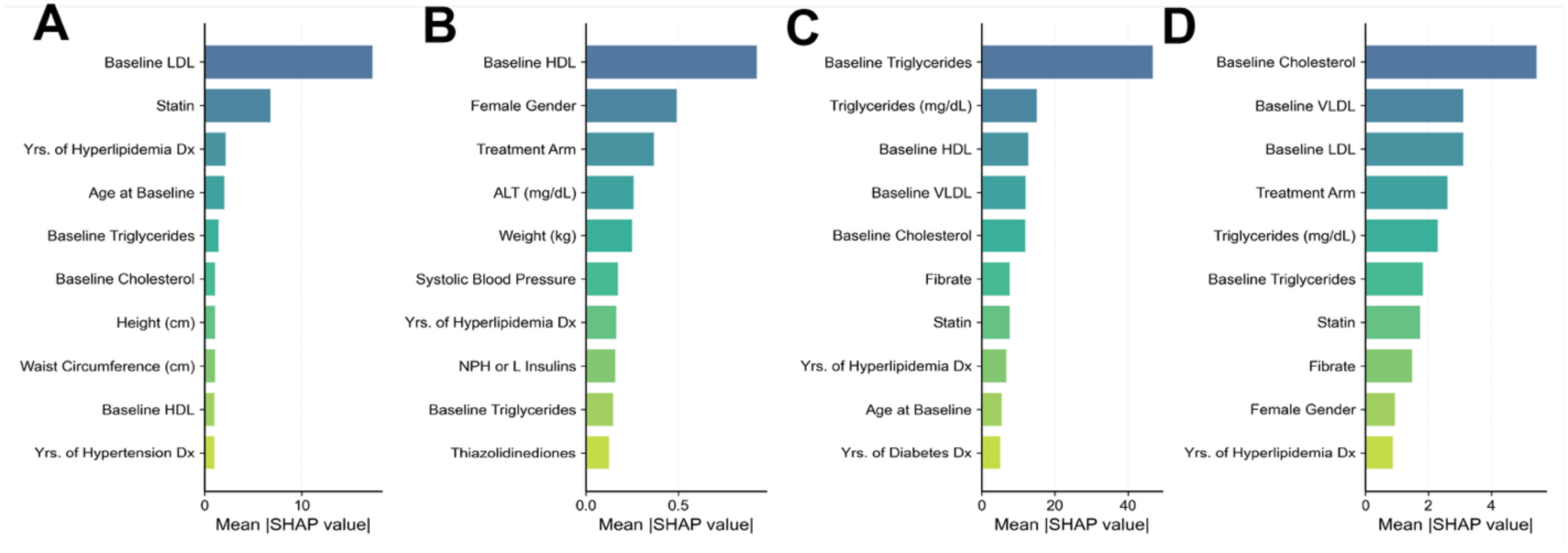
Feature importance analysis. Plotted are SHAP importances for **(A)** LDL-C, **(B)** HDL-C, **(C)** TG, and **(D)** VLDL, reflecting the mean SHAP value for each of the 10 most important variables for each prediction paradigm.

### Uncertainty Quantification via Conformal Prediction

To translate point predictions into clinically actionable intervals, we applied conformal prediction using 𝛼 = 0.1 (i.e., specified coverage of 90%). We validated the calibration of these intervals by comparing the target coverage probability (1 − 𝛼) against the empirical coverage (the actual percentage of true values falling within the predicted interval) on the test set. Across all lipid fractions, empirical coverage closely matched nominal levels, confirming that the intervals were neither overly conservative nor prone to undercoverage. The illustrative example in **Figure 4** highlights how the method adapts interval width to individual patient uncertainty: for LDL-C, where model predictions exhibited moderately high fidelity, the intervals remained relatively narrow, whereas triglyceride predictions, characterized by substantial biological volatility, led to markedly wider intervals.

**Figure 4.**
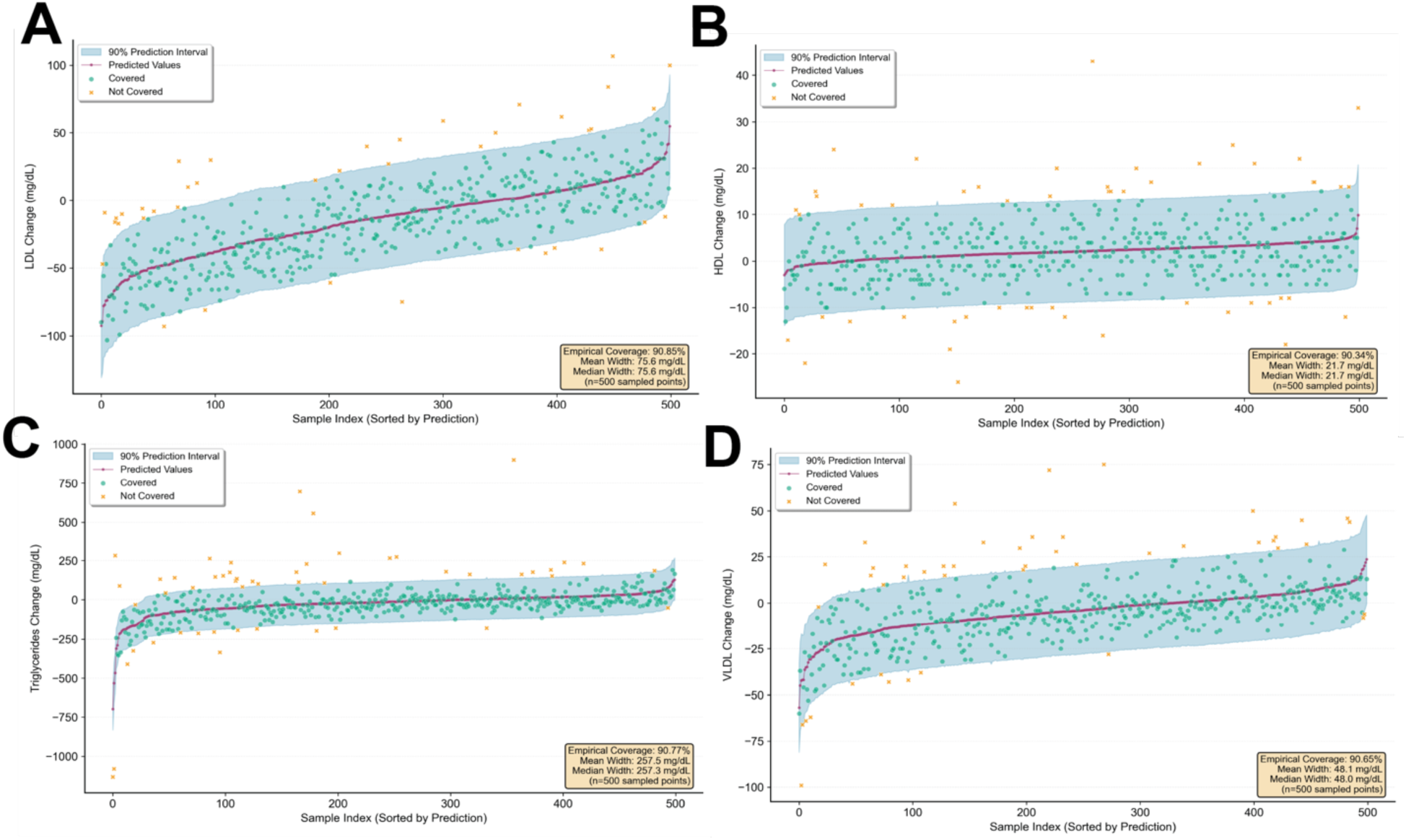
Example results for the conformal prediction task. Example conformal prediction plots for **(A)** LDL-C, **(B)** HDL-C, **(C)** TG, and **(D)** VLDL, sorted by predicted value. Predicted values are shown via the purpose line, the 1 − 𝛼 prediction interval is indicated by blue shading, and ground truth points that are covered within the prediction interval (green) or not covered (yellow) are shown via colored markers.

As detailed in **Table 2**, the model achieved excellent calibration across all lipid types. For LDL-C, the 90% prediction intervals (𝛼 = 0.10) achieved an empirical coverage of 90.9%, with a mean interval width of 75.6 mg/dL. This implies that for a given patient, the model can bound the expected LDL change within a range of approximately ± 38 mg/dL with 90% confidence.

The difficulty in predicting HDL-C was reflected in the uncertainty metrics; while coverage remained accurate (90.3% for 𝛼 = 0.10), the intervals were wide relative to the total dynamic range of HDL changes. Conversely, for Triglycerides, the 90% confidence intervals spanned an average of 257.5 mg/dL, underscoring the high volatility of this metric in a diabetic population.

To assess whether conformal prediction intervals maintained valid coverage across the full range of predicted lipid changes, we examined empirical coverage within each decile of predicted values (**Figure 5**). While overall coverage closely matched the nominal 90% target for all lipid fractions, a consistent pattern of undercoverage emerged in the lowest prediction bins. For the first decile, which corresponds to patients with the smallest predicted lipid changes, empirical coverage fell substantially below the 90% threshold. This undercoverage was most pronounced for triglycerides and VLDL, where the lowest bins missed the target by over 15 percentage points. In contrast, coverage for middle and upper prediction bins consistently met or exceeded the nominal rate, often ranging from 90-96%. This systematic pattern suggests that the model’s uncertainty estimates are less reliable for patients predicted to have minimal lipid responses, potentially reflecting greater residual heterogeneity or unmeasured confounding among individuals near their physiological set points. Clinicians should interpret prediction intervals with additional caution for patients expected to show modest treatment effects, while intervals for larger predicted changes appear well-calibrated.

**Figure 5.**
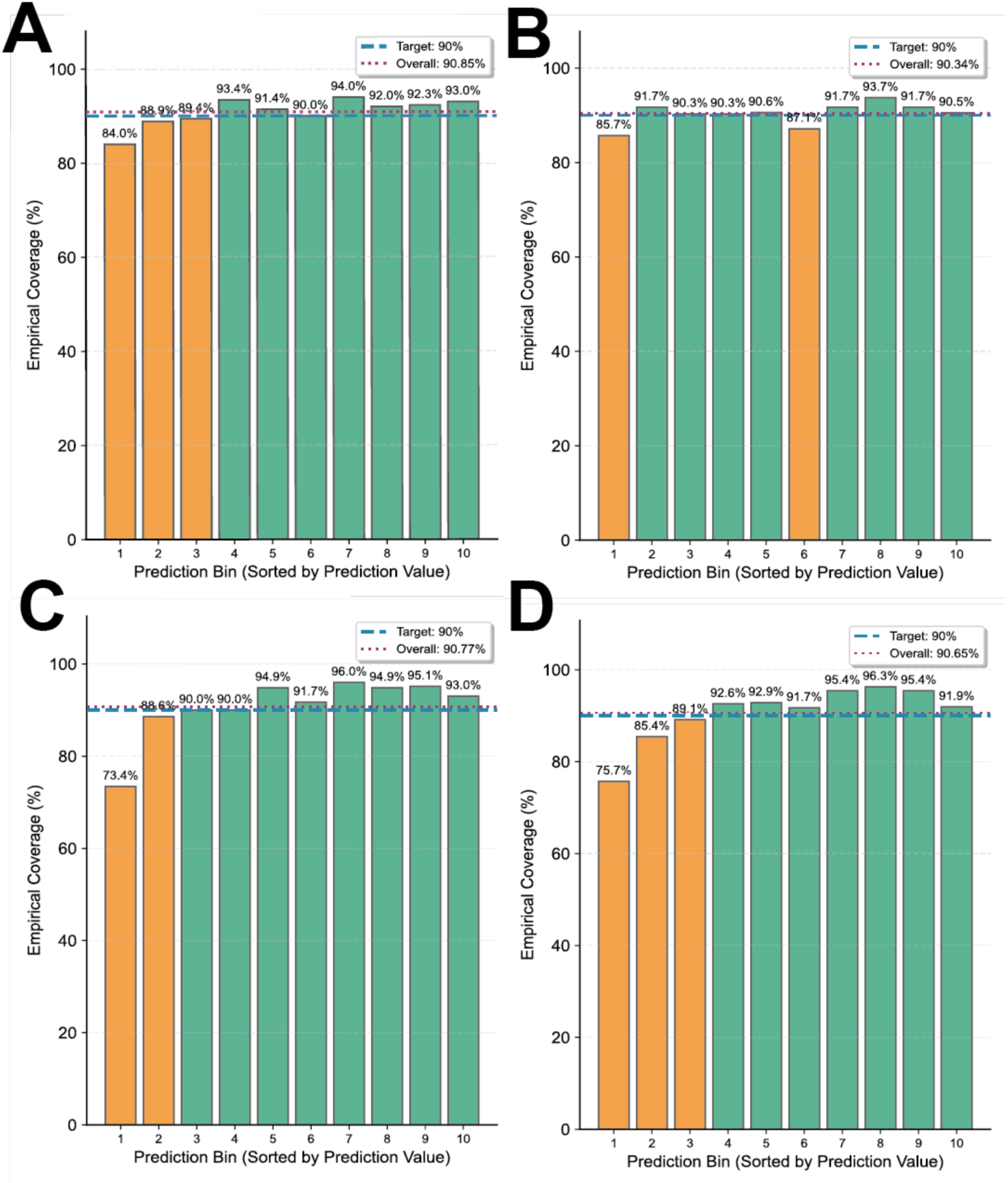
Coverage by prediction bin. Plotted is the empirical coverage for **(A)** LDL-C, **(B)** HDL-C, **(C)** TG, and **(D)** VLDL. Coverage is divided by prediction bin (i.e., each decile of predicted value), with target coverage (90%; blue dotted line) and overall empirical coverage (dotted red line) overlayed.

Taken together, these results demonstrate that conformal prediction successfully provides calibrated uncertainty estimates for individualized lipid response prediction. Across all lipid fractions, overall empirical coverage closely matched the specified 90% confidence level, confirming that the distribution-free guarantees of conformal prediction hold in this clinical context. The prediction intervals appropriately scaled to reflect inherent biological variability, being narrower for more predictable outcomes like LDL-C and substantially wider for volatile fractions like triglycerides, thus providing clinicians with an intuitive measure of prediction reliability. While coverage was attenuated for the lowest prediction bins, the framework nonetheless offers a principled approach to communicating model uncertainty that moves beyond point estimates toward actionable confidence ranges suitable for clinical decision support.

## Discussion

In this analysis of the ACCORD trial cohort, we applied ML techniques to quantify and predict longitudinal lipid responses in patients with T2DM. Our findings reveal a distinct hierarchy of predictability: LDL-C changes are moderately predictable using standard clinical variables (𝑅^2^≈ 0.50), whereas HDL-C trajectories are largely stochastic relative to the available data (𝑅^2^ ≈ 0.06). Furthermore, by employing conformal prediction, we demonstrated that it is possible to generate rigorous, coverage-guaranteed uncertainty intervals for these predictions. This study highlights both the potential and the limitations of EHR phenotypes in guiding precision lipid management.

### Heterogeneity of Lipid Response in Type 2 Diabetes

The primary finding that LDL-C response is substantially more predictable than HDL-C or TG response likely reflects the distinct biological mechanisms governing these lipoproteins. Statin therapy, the dominant intervention in this cohort, has a potent, direct mechanism of action (HMG-CoA reductase inhibition^35^) that linearly correlates with dose and baseline LDL-C burden. Our feature importance analysis confirmed that baseline LDL-C and statin usage were the dominant predictors, consistent with the “law of initial values” where patients with higher baselines exhibit the steepest absolute drops.

In contrast, the poor predictive performance for HDL-C (𝑅^2^ < 0.10) suggests that HDL-C modulation in T2DM is driven by factors largely uncaptured in standard clinical trial datasets. This aligns with broader literature suggesting that HDL-C levels are heavily influenced by germline genetics^36,37^, detailed lifestyle factors (e.g., specific dietary composition, exercise intensity)^38–40^, and alcohol consumption^41–43^: granular variables often missing or poorly quantified in clinical datasets. Consequently, while clinicians can confidently anticipate LDL-C reductions, HDL-C responses remain elusive targets for prediction.

The move from population-level guidelines to personalized titration is the central goal of precision medicine. Our results suggest that ML models can serve as “guardrails” for LDL-C management. For example, a model predicting a modest LDL-C reduction for a specific patient, despite high-intensity statin prescription, could alert clinicians to potential non-responder status early. This could prompt earlier initiation of adjunct therapies (e.g., ezetimibe^44^ or PCSK9 inhibitors^45,46^) rather than waiting for a follow-up visit to confirm failure. However, the moderate 𝑅^2^ values (ranging from 0.30 to 0.50 for non-HDL-C fractions) indicate that a “lipidome-blind” approach, which relies solely on clinical history and basic labs, hits a performance ceiling. As noted in our introduction, the missing variance is likely hidden within the gut microbiome and genetic polymorphisms. Future predictive frameworks must integrate polygenic risk scores and metabolomic data to bridge the gap between our model’s current performance and the near-perfect prediction required for fully automated dosing.

A critical contribution of this work is the application of conformal prediction to clinical decision support. Standard regression models provide point estimates can be misleadingly precise. In medicine, the cost of error is high. By generating calibration-guaranteed prediction intervals (e.g., “Your LDL-C will drop by 15–45 mg/dL with 90% confidence”), we provide clinicians with a measure of reliability. Our results showed that these intervals vary significantly by patient and lipid type. For variables like TG, where the 90% confidence interval spanned over 250 mg/dL, the model effectively signals to the clinician that the outcome is volatile and that close monitoring is required.

The strengths of this study include the use of a large, well-phenotyped multi-center cohort and the implementation of robust validation techniques (conformal prediction). However, several limitations must be noted. First, the ACCORD trial data, while rich, reflects therapeutic practices from a specific era; the impact of modern agents like SGLT2 inhibitors or GLP-1 receptor agonists on lipid predictability was not assessed^47^. Second, adherence relies on self-reported metrics, which are imperfect proxies; unmeasured non-adherence is a major source of noise in treatment response modeling. Finally, our model implies association, not causality; while feature importance highlights predictive markers, these may represent confounding comorbidities rather than mechanistic drivers of lipid change.

Looking forward, several avenues may substantially enhance the precision and clinical utility of individualized lipid-response modeling. Integrating richer biological data, such as polygenic risk scores, targeted metabolomics, and gut microbiome signatures, may help capture the variance underlying the unpredictable components of HDL-C and triglyceride response, particularly in patients with type 2 diabetes. Combining these data streams with modern cardiometabolic therapies will also be essential to ensure model relevance in contemporary practice. Methodologically, extending conformal prediction to dynamic, time-updated models and exploring causal inference frameworks may yield more robust estimates of patient-specific treatment effects and further mitigate risks associated with black-box predictions. Finally, prospective evaluation of these methods in real-world clinical workflows will be necessary to determine whether algorithmically derived predictions, as well as their associated uncertainty intervals, meaningfully improve therapeutic decision-making and reduce residual cardiovascular risk.

### Conclusion

We demonstrate that machine learning models can predict LDL-C and TC responses with clinically relevant accuracy in patients with T2DM, while HDL-C response remains largely unpredictable using standard clinical features. The integration of conformal prediction provides a necessary safety layer for AI-driven clinical tools, translating abstract error metrics into actionable confidence intervals. These findings support the development of algorithmic decision aids for lipid management, provided they are deployed with transparent uncertainty quantification and an understanding of the biological limits of prediction using EHR data alone.

## Data Availability

Data from the ACCORD study is available upon request to the National Heart, Lung, and Blood Institute.

https://biolincc.nhlbi.nih.gov/studies/accord/

## Acknowledgements

This material is based upon work supported by the National Science Foundation Graduate Research Fellowship under Grant No. DGE2139757, awarded to CH. Any opinion, findings, and conclusions or recommendations expressed in this material are those of the authors(s) and do not necessarily reflect the views of the National Science Foundation.

## Code and data availability statement

Code will be released as a public GitHub repository upon acceptance of the manuscript. As a proprietary dataset, ACCORD data cannot be released by the authors but is available by request to the National Heart, Lung, and Blood Institute.

## Contributions

P.S. and C.H. conceived of the project. C.H. conducted analyses. C.H. and D.O. wrote the paper, with advisement from P.S. and R.C. All authors reviewed and approved the final draft.

## Correspondence and request for materials

Should be addressed to C.H.

## Supplementary Tables

**Table S1.** Comparison of prediction performance across candidate ML models. Bolded values indicate the highest performing model, and underlined values indicate the second-highest performing.

| Lipid fraction | Model | $R^2$ | MAE |
| --- | --- | --- | --- |
| LDL | Lasso | **0.519** | **18.32** |
|  | Ridge | <u>0.516</u> | <u>18.42</u> |
|  | XGBoost | 0.505 | 18.71 |
|  | Random forests | 0.500 | 18.73 |
|  | ElasticNet | 0.411 | 20.36 |
| HDL | Lasso | 0.041 | 5.14 |
|  | Ridge | <u>0.044</u> | 5.14 |
|  | XGBoost | **0.049** | **5.10** |
|  | Random forests | 0.044 | <u>5.12</u> |
|  | ElasticNet | < 0 | 5.56 |
| Triglycerides | Lasso | **0.187** | <u>64.08</u> |
|  | Ridge | <u>0.182</u> | 64.93 |
|  | XGBoost | 0.134 | 65.69 |
|  | Random forests | 0.179 | **63.35** |
|  | ElasticNet | 0.120 | 66.87 |
| VLDL | Lasso | **0.169** | <u>11.79</u> |
|  | Ridge | <u>0.169</u> | 11.86 |
|  | XGBoost | 0.126 | 12.12 |
|  | Random forests | 0.168 | **11.75** |
|  | ElasticNet | < 0 | 12.78 |
| Total cholesterol | Lasso | **0.411** | <u>23.19</u> |
|  | Ridge | <u>0.408</u> | **23.07** |
|  | XGBoost | 0.385 | 23.88 |
|  | Random forests | 0.398 | 23.70 |
|  | ElasticNet | 0.305 | 25.36 |

**Table S2.** Hyperparameter selection. Shown are candidate parameter-value pairs for hyperparameters considered across the five candidate models.

| **Model** | **Hyperparameter** | **Parameter set** |
| --- | --- | --- |
| Lasso | alpha | [0.001, 0.01, 0.1, 1, 10, 100] |
|  | max_iter | [1000, 2000, 3000] |
| Ridge | alpha | [0.01, 0.1, 1, 10, 100] |
|  | Solver | ['auto', 'svd', 'cholesky', 'lsqr', 'sparse_cg', 'sag', 'saga'] |
| XGBoost | n_estimators | [50, 100, 200] |
|  | max_depth | [3, 4, 5] |
|  | learning_rate | [0.01, 0.1, 0.2] |
|  | subsample | [0.7, 0.8, 0.9] |
| RF | n_estimators | [50, 100, 200, 300] |
|  | max_depth | [3, 5, 7, 10, None] |
|  | min_samples_split | [2, 5, 10] |
|  | min_samples_leaf | [1, 2, 4] |
|  | max_features | ['sqrt', 'log2', None] |
| ElasticNet | alpha | [0.001, 0.01, 0.1, 1, 10, 100] |
|  | l1_ratio | [0.0, 0.25, 0.5, 0.75, 1.0] |

**Table S3.** Conformal prediction results. Shown is the empirical coverage and mean interval width (and standard deviation of width) for the LASSO model evaluated on the range 𝛼 = 0.01, 0.05, 0.10, 0.20 across the five lipid categories considered.

| **Prediction** | **Alpha**<br>**(coverage)** | **Empirical coverage**<br>**(%)** | **Mean width (mg/dL)**<br>**± STD** |
| --- | --- | --- | --- |
| **LDL** | 0.01 (99%) | 99.1 | 143.0 ± 0.6 |
|  | 0.05 (95%) | 95.5 | 94.1 ± 0.3 |
|  | 0.10 (90%) | 90.9 | 75.6 ± 0.2 |
|  | 0.20 (80%) | 80.7 | 56.8 ± 0.2 |
| **HDL** | 0.01 (99%) | 99.1 | 43.6 ± 0.3 |
|  | 0.05 (95%) | 95.7 | 27.4 ± 0.2 |
|  | 0.10 (90%) | 90.3 | 21.6 ± 0.2 |
|  | 0.20 (80%) | 82.2 | 15.9 ± 0.1 |
| **Triglycerides** | 0.01 (99%) | 99.5 | 796.6 ± 33.4 |
|  | 0.05 (95%) | 96.3 | 359.3 ± 38.2 |
|  | 0.10 (90%) | 90.8 | 251.5 ± 39.2 |
|  | 0.20 (80%) | 81.8 | 174.3 ± 6.0 |
| **VLDL** | 0.01 (99%) | 99.1 | 122.0 ± 3.9 |
|  | 0.05 (95%) | 95.6 | 63.9 ± 4.3 |
|  | 0.10 (90%) | 90.7 | 47.6 ± 4.3 |
|  | 0.20 (80%) | 80.8 | 34.2 ± 0.9 |
| **Total**<br>**cholesterol** | 0.01 (99%) | 99.8 | 194.8 ± 9.0 |
|  | 0.05 (95%) | 98.0 | 117.6 ± 9.5 |
|  | 0.10 (90%) | 90.5 | 93.2 ± 9.3 |
|  | 0.20 (80%) | 88.0 | 70.5 ± 1.4 |

## Supplementary Figures

**Figure S4.**
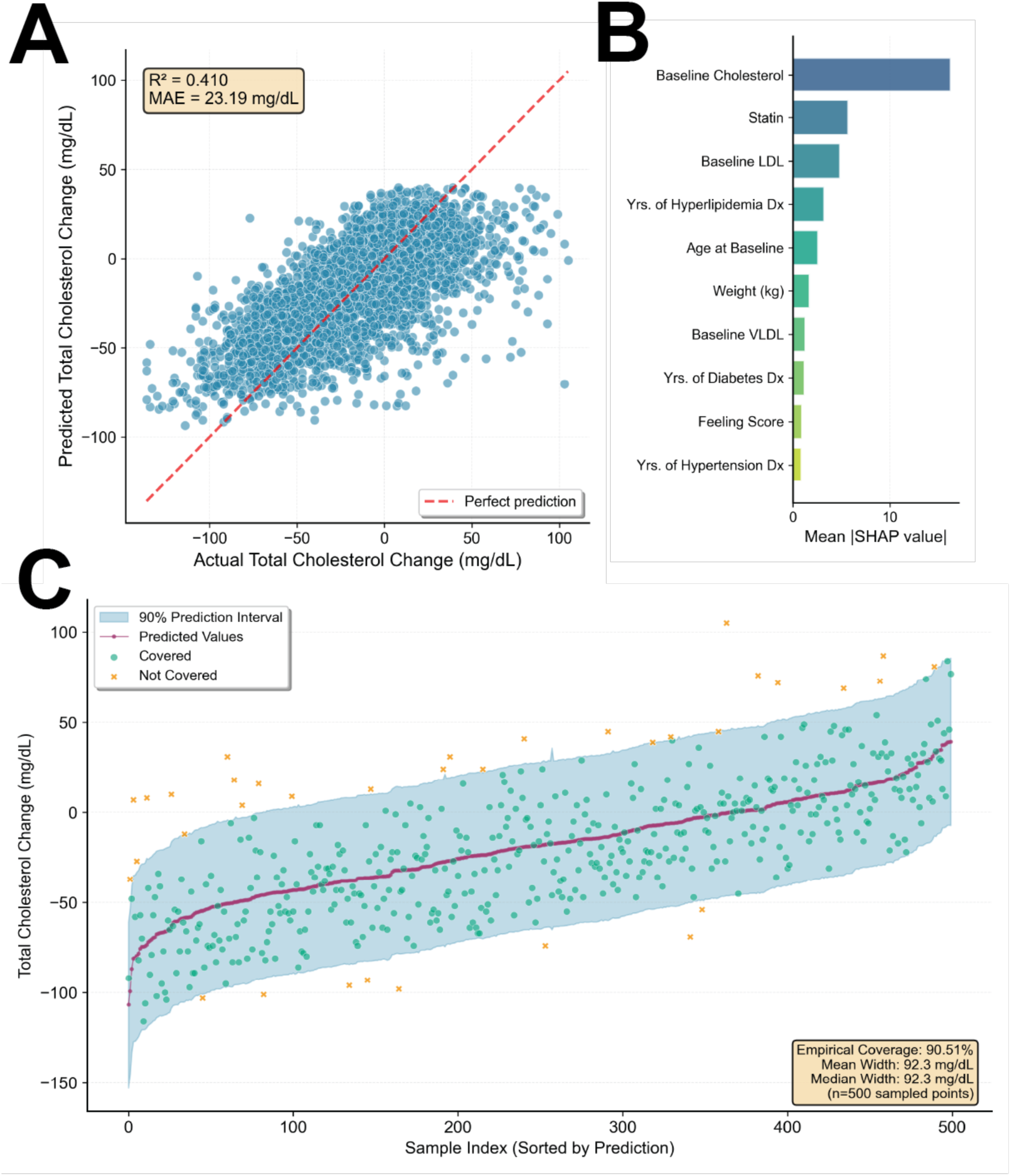
Prediction results for total cholesterol. Plotted is **(A)** actual vs. predicted values, **(B)** SHAP feature importance, and **(C)** conformal prediction results for TC.

